# Pediatric nurses’ perspective on the culture of patient safety

**DOI:** 10.1101/2025.06.08.25329208

**Authors:** Zolikha Mohseni, Mahshid Mirzaei, Zahra Taheri Ezbarami, Ehsan Kazemnejad Leili

## Abstract

**Background:** Patient safety has been introduced as an epidemic concern, and worldwide, many patients, including children, are affected by medical errors that endanger their safety. This study aimed to patient safety culture from the perspective of pediatric nurses and related factors.

**Design and Methods:** In this analytical-cross-sectional study, that was adopted between April 2023 and September 2023, 200 nurses worked in the pediatrics’ wards using the census sampling method. The instruments used were a demographic and a patient safety culture questionnaire. Data were analyzed using Chi-2, Fisher inferential tests, and regression analysis in SPSS software version 24.

**Results:** Nurses’ patient safety culture was in the moderate range (55.3 ± 16.9), and nurses with older age, formal employment, fixed work shifts, higher work experience, having overtime, participating in training workshops, and not having Stress during the work shift had a significantly higher safety culture (P<0.05). “Intra-departmental teamwork” had the highest score, and “non-punitive response when errors” had the lowest score for patient safety culture.

**Conclusion:** Based on the findings, the safety culture among nurses working in children’s wards is not favorable, so it is suggested to design training programs and create a safe environment to track errors in hospitals.

**Clinical evidence:** The “non-punitive response in case of error” dimension has the lowest score in the cultural dimensions of patient safety. There are problematic areas in the safety culture of the sector, which shows that this culture needs to be better developed, with special attention to the dimensions of the culture that presented a less positive evaluation

- **What is currently known?**

The culture of patient safety is the primary and necessary component for promoting and improving patient safety, and children, as a high-risk group, are under the threat of non-compliance with patient safety. The patient safety culture in children’s ward nurses is unfavorable.

- **And What does this article add?**

The “non-punitive response in case of error” dimension has the lowest score in the cultural dimensions of patient safety. There are problematic areas in the safety culture of the sector, which shows that this culture needs to be better developed, with special attention to the dimensions of the culture that presented a less positive evaluation

## Introduction

One of the most basic and essential requirements for health and medical services is not harming the patient while providing care (1). Patient safety in the health service system of all countries means compliance with principles that, if applied correctly, will not harm the client or minimize potential harm (2). The American Institute of Medicine has considered patient safety as the most crucial factor for providing quality care, which is the prevention of possible injuries intentionally or unintentionally caused to the patient by people and the medical system (3). All over the world, one out of every ten patients is affected by medical errors, and this has caused the World Health Organization to introduce patient safety as an epidemic concern (4). Observing and maintaining the safety of the patients is one of the professional and ethical duties of all health service providers (5).

Meanwhile, compared to adults, hospitalized children are three times more exposed to damage caused by errors (6). Their safety is more endangered (7), and although compliance with patient safety tips is mandatory in all parts of the hospital and leads It is possible to reduce risks. However, it is more complicated in children’s departments due to the unstable anatomical and physiological characteristics of children, who are very fragile and vulnerable (4, 8), so research results show that about 30% of children have at least one About 5% of them experience being hospitalized several times during their childhood (9). Among them, 3-17% suffer injuries or complications due to medical errors (10). In England, more than one-third of deaths are due to improper management of patients related to safety. Also, more than 16% of patients have experienced an injury caused by non-observance of safety at least once (11, 12).

Among these, the principal and essential component of healthcare provider organizations to promote and improve patient safety is the patient safety culture (4). Currently, most of the developed countries have realized that simply having management systems and advanced technology is not enough to achieve sustainable development but promoting safe behaviors in employees, their values, beliefs, and attitudes towards safety, as well as the organization’s attitude towards safety, which Basically, the culture forms their safety, it is the way to prevent mistakes (13). The patient safety culture shows the priority level of patient safety from the point of view of employees in the department and their workplace organization(14). This term was used for the first time in 1986 after the Chornobyl accident in Russia, and special attention to safety culture was emphasized in different countries (15). Creating a positive safety culture will allow healthcare providers to discuss, analyze and report errors and near-error cases, which is a big step in improving the quality of healthcare for children (4).

Due to the increase in medical and nursing errors, it is essential to be aware of the patient safety culture in order to change this culture and make it compatible with the advances made in the field of care quality because improving patient safety is not only a clinical issue but also related to organizational dimensions (6). A study conducted in two specialized children’s hospitals in Tehran regarding the rate of medication errors and self-reported obstacles by nurses found that 76% of nurses had medication errors in the past year. However, only 28% of them reported their errors, and the most common reason for this behavior was the fear of the impact of the error on the patient, giving compensation, worrying about the lousy record after reporting, the impact on the annual evaluation, and not being supported by managers (16).

Nurses constitute the largest group of healthcare workers whose primary responsibility, especially pediatric nurses, is to provide safe nursing services to children and families. Based on the defined duties of the nurse, relying on the charter of nursing rights based on the safety process and care standards, it is the responsibility of the nurse to try and follow up to plan and implement nursing measures in order to protect the child from injuries caused by accidents and physical risks during hospitalization (7). Their point of view is essential for patient safety, and their participation will improve the health system and patient safety(6).

Error in the care of patients is a non-independent concept; that is, it cannot be said that only nurses in pediatric wards in Iran make errors, but it can be said that nurses’ understanding of the concept of error in pediatric wards is different from each other, and this can create many risks for children and hospitalized patients (17). Because nurses are usually the first line of defense to maintain safety in the children’s department, the safety culture prevailing in the hospital may affect nurses’ performance. Therefore, it is necessary to take a different look at the safety culture of nurses working in the children’s department by stating the existing weaknesses and strengths and the necessary solutions to improve the safety and culture of the nurses in the children’s department; the present study aims to determine the patient safety culture. This study studied patient safety culture from the perspective of pediatric nurses and related factors.

## Methods

In this descriptive-analytical cross-sectional study, a total of 200 pediatrics nurses, by complete count sampling method, participated. The inclusion criteria were as follows: Consent to participate in the research and to have at least six months of work experience in pediatrics departments.

The data collection tool in this study included two sections. The first part includes a person’s demographic information with Occupational factors (work experience, work experience in the children’s department, type of employment, name of the department, type of work shift, number of patients under observation, time of care, number of nurses in the desired shift, overtime), individual factors (age, sex, education degree, Marital status, having children, number of children, monthly salary, having stress during shift) and organizational factors (organizational position, promotion history, possibility of promotion in the organization, familiarity with the description of duties in the organization).

The second part is the Hospital Survey on Patient Safety Culture Questionnaire (Hsopsc). This questionnaire, which is designed to assess the level of Patient Safety Culture in nurses, has 42 questions in 12 different dimensions of patient safety (error reports, general understanding of patient safety, organizational learning and continuous improvement, supervisor’s actions regarding safety promotion, intra-departmental teamwork, informing employees of errors and giving feedback, clear communication, The non-punitive response in case of errors is the medical staff and their related issues, the support of the hospital management for the patient, the transfer of the patient and the exchange of information related to him, the number of reported errors). The minimum score that can be obtained from this questionnaire is 1 and the maximum score is 5, and higher scores mean a higher safety culture, and there are three levels of safety culture: high (more than 75% of answers agree), medium (between 50-75% of answers) agree) and low safety culture (less than 50% of answers agree) were classified. This questionnaire’ Cronbach’s alpha was evaluated by Mughri et al. and was 0.82 and the Spear-Brown coefficient was 0.81 (21). the reliability of the questionnaire was also investigated by conducting a preliminary study on 25 nurses and using the test-retest method, and Cronbach’s alpha values were estimated at 0.72.

Data collection was provided to the nurses in the morning and evening shifts and then collected. The collected data were analyzed by descriptive and inferential statistical tests under SPSS version 21 software. Spearman’s correlation coefficient was used to determine the relationship between factors related to safety culture.

## Results

In general, this questionnaire was completed by 200 qualified nurses who met the study entry criteria. All nurses were women (100 percent) with an average age of 36.7 ± 7.6 years. The majority of these nurses were married (81%) and had children (64%), and the majority had a total work experience of sixteen years or more (32.5%), and 58.5% of them had a work experience of less than 5 years in the children’s department. 41% of the participating nurses worked overtime between 30 and 60 hours with an average of 23.9 ± 1.34 hours per month and 58% stated that they have enough time to perform their duties. The majority of nurses (83.5%) participated in a training workshop related to patient safety, were interested in the nursing profession (68%) and did not have a second job (81%) (Table 1).

**Table 1:**
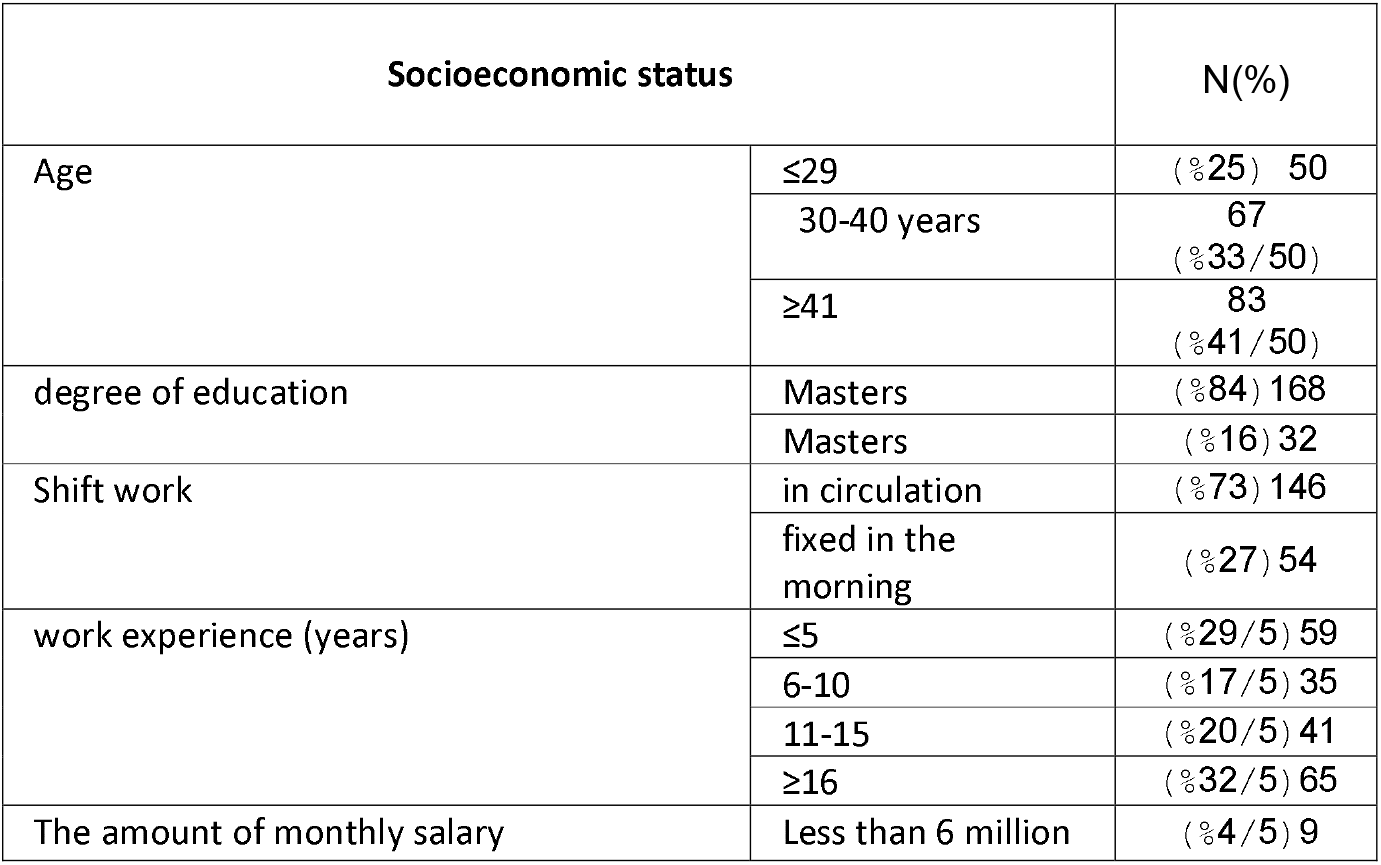
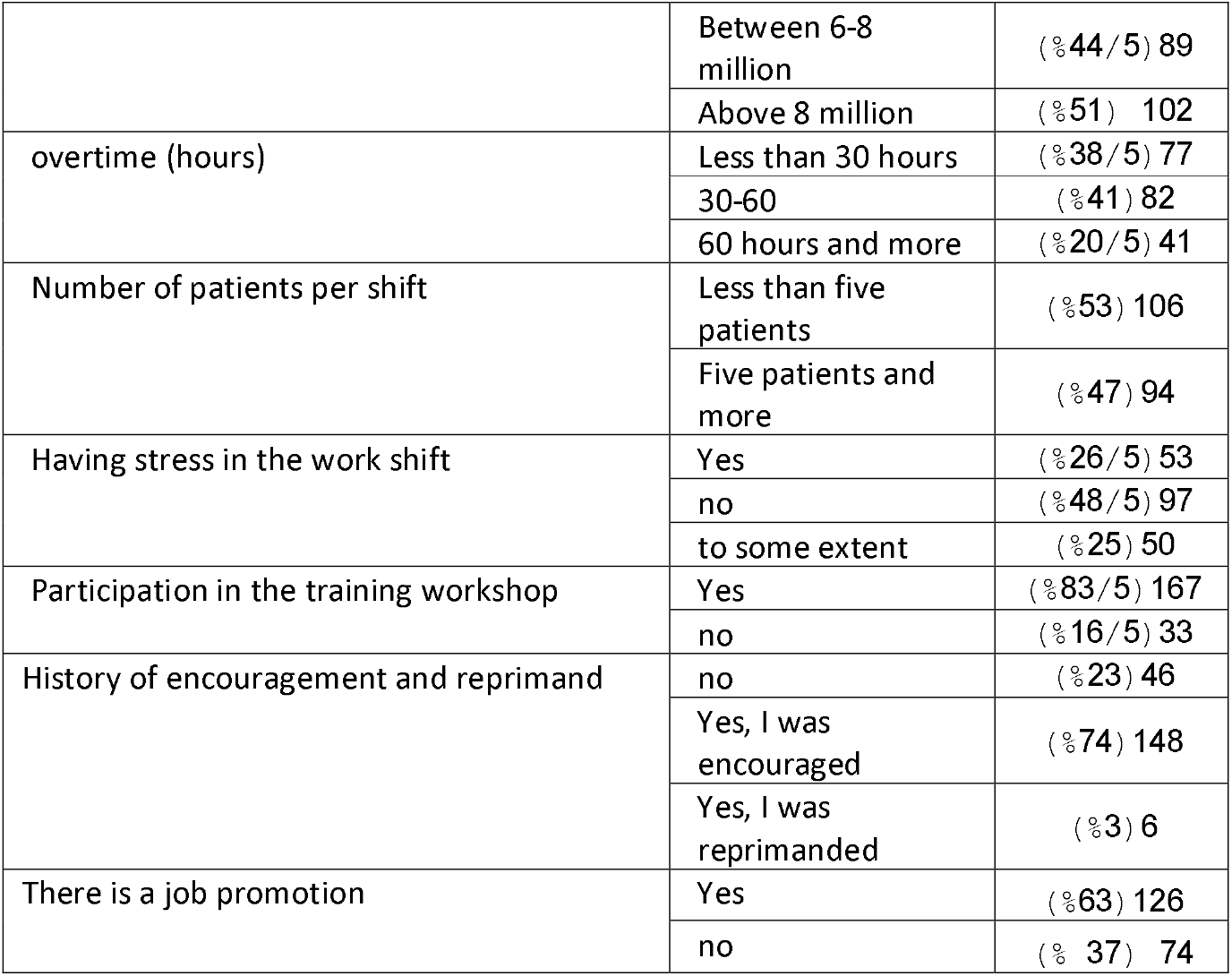
Socioeconomic status of the sample.

The mean and standard deviation of the nurses’ safety culture score was 55.3 ± 16.9, and the status of patient safety culture was “low” in 37% of nurses, and “high” in only 17% of them. The highest average score of patient safety culture is related to the dimension of “intra-departmental teamwork” with a mean and standard deviation of 78.6 ± 21.7 and the lowest average score is related to the dimension of “non-punitive response in case of errors” with a mean and standard deviation of 2. It was 21.2 ± 28. Also, the patient safety culture was significantly influenced by the factors of age group, having children and having stress in the work shift, service department, type of employment, rotating work shift, work experience, overtime, having a second job and nurses who are encouraged in the organization. (P<0.05) (Table 2).

**Table 2:** Comparing patient safety culture according to socioeconomic-occupational factors.

| Statistical index<br>socioeconomic -occupational factors |  | patient safety culture |  |  |  |  |  | P-Value |
| --- | --- | --- | --- | --- | --- | --- | --- | --- |
|  |  | low |  | medium |  | high |  |  |
|  |  | N | % | N | % | N | % |  |
| Age | less than 30 years | 17 | 34 | 28 | 56 | 5 | 10 | *0/029 |
|  | 40-30 | 30 | 44/8 | 30 | 44/8 | 7 | 10/4 |  |
|  | 40 years and more | 27 | 32/5 | 34 | 41 | 22 | 26/5 |  |
| degree of education | Masters | 58 | 34/5 | 79 | 47 | 31 | 18.5 | *0/194 |
|  | Masters | 16 | 50 | 13 | 40/6 | 3 | 9/4 |  |
| Shift work | in circulation | 60 | 41/1 | 69 | 47/3 | 17 | 11/6 | *0/003 |
|  | fixed in the morning | 14 | 25/9 | 23 | 42/6 | 17 | 31/5 |  |
| work experience | Five years and less | 21 | 35/6 | 32 | 54/2 | 6 | 10/2 | *0/003 |
|  | 11-6 | 21 | 60 | 13 | 37/1 | 1 | 2/9 |  |
|  | 11-16 | 13 | 31/7 | 20 | 48/8 | 8 | 19/5 |  |
|  | 16 years and older | 19 | 29/2 | 27 | 41/5 | 19 | 29/2 |  |
| Overtime | Less than 30 hours | 19 | 24/7 | 31 | 40/3 | 27 | 35/1 | <0/001 |
|  | 60-30 | 31 | 37/8 | 44 | 52/7 | 7 | 8/5 |  |
|  | 60 hours and more | 24 | 58/5 | 17 | 41/5 | 0 | 0 |  |
| Number of patients per shift | Less than five patients | 40 | 37/7 | 43 | 40/6 | 23 | 21/7 | *0/110 |
|  | Five patients and more | 34 | 36/2 | 49 | 52/1 | 11 | 11/7 |  |
| Having stress in the work shift | Yes | 30 | 56/6 | 18 | 34 | 5 | 9/4 | *0/006 |
|  | no | 26 | 26/8 | 49 | 50/5 | 22 | 22/7 |  |
|  | somewhat | 18 | 36 | 25 | 50 | 7 | 14 |  |
| Participation in the training workshop | Yes | 56 | 33/5 | 80 | 47/9 | 31 | 18/6 | *0/063 |
|  | no | 18 | 54/5 | 12 | 36/4 | 3 | 9/1 |  |
| History of encouragement and reprimand | no | 19 | 41/3 | 24 | 52/2 | 3 | 6/5 | **0/013 |
|  | Yes, I was encouraged | 50 | 33/8 | 68 | 45/9 | 30 | 20/3 |  |
|  | Yes, I was reprimanded | 5 | 83/3 | 0 | 0 | 1 | 16/7 |  |
| There is a job promotion | Yes | 42 | 33/3 | 58 | 46 | 26 | 20/6 | *0/143 |
|  | no | 32 | 43/2 | 34 | 45/9 | 8 | 10/8 |  |
| Access to safety resources in the workplace | Yes | 63 | 36/2 | 81 | 46/6 | 30 | 17/2 | *0/834 |
|  | no | 11 | 42/3 | 11 | 42/3 | 4 | 15/4 |  |

\*Chi-square test \*\*Fisher's exact test

## Discussion

To the best of our knowledge, this study is the first to Predict the culture of patient safety from the perspective of pediatric nurses.

The results of this study show that the patient safety culture of nurses working in children’s wards was in the medium range, and the majority of nurses generally reported the status of patient safety culture as medium or low. Only 35% of the nurses working in the children’s department in the study by Macedo et al. (2016) reported the patient safety culture in an excellent range (35%) (6). Obtaining average and poor results in the patient safety culture in the studies of Tamazoni et al. (2015) among nurses working in the neonatal ward has also been confirmed that the majority of nurses report the safety status of patients at an average level (18), Arshaddi Bostan Abad et al. 2015) after examining the patient safety culture in neonatal intensive care units, which reports that the scores obtained in 12 cultural dimensions of patient safety from the nurses’ point of view were in the average range, and the level of patient safety culture in neonatal intensive care units was within It is low and medium (4). Al-Ahmadi et al. (2010) also reported the average patient safety culture in the medium range (19). The results of this section are consistent with the score obtained from the sum of the scores in the field of general understanding of patient safety in the studies of Mohammadi et al. (2020) (20), Beqaei et al. This is even though in the studies conducted in America and Turkey on nurses working in public departments, 74% and 42% of the studied people, respectively, considered the safety status of their work unit to be very good and excellent (21-23).

In the present study, the dimension of “intra-departmental teamwork” received a high safety culture score. In terms of the high status of the patient’s safety culture, it also received a high percentage. In line with this result, Macedo et al. (2016), after examining the patient safety culture from the point of view of nurses working in children’s wards, admit that teamwork (62%) is one of the dimensions that has received the highest number of positive responses (6). Although in the studies conducted by Zwijnenberg et al. (2016) in America (22), Top et al. (2015) in Turkey (21), Stittelaar et al. (2016) in Taiwan (24), and Thomas et al. (2016) (25) In the Netherlands, the dimension of teamwork within the hospital units has obtained the highest score, which is in line with the results of the present study. In the study by Wang et al. (2014) conducted in seven public hospitals on 463 nurses, 87% of A positive evaluation was assigned to the dimension of teamwork (26). However, Salvati et al. (2013), in a study examining the culture of patient safety from the perspective of nurses, admitted that teamwork among hospital units has the least number of positive responses. Hence, the nurses believed there needed to be better coordination between hospital units and often faced problems when coordinating and exchanging information with other hospital units (27).

Obtaining the present results that the nurses assigned the highest score to the teamwork dimension was not far from expected because group and teamwork are the strength of the nursing profession (28). Nursing is a profession that has the nature of doing work in a team and cooperative manner. There is a lot between employees, and it is only possible to take care of the patient with cooperation with other medical staff members. Therefore, nurses in the culture of patient safety also rely on cooperation with each other, and teamwork is essential because of their suggestions.

In the present study, “organizational learning and continuous improvement” received the highest score for safety culture, and in terms of the high status of patient safety culture, it also received a high percentage. In line with this result, Macedo et al. (2016) after examining the patient safety culture from the view of nurses working in children’s wards, admit that organizational learning and continuous improvement (58%) are among the dimensions that have received the highest number of positive responses(6). and in the studies of Mohammadi et al. (2020) and Shafiei et al. (2014), organizational learning and continuous improvement have the most positive responses among the two dimensions examined (20, 26).

Obtaining a high score in the dimension of organizational learning and continuous improvement shows the continuation of the implementation of learning programs and continuous improvement of security in children’s departments. The results of the present study show the important point that from the point of view of nurses, education and efforts to improve the level of knowledge can lead to improving the cultural status of patient safety.

The dimensions of “non-punitive response in case of errors” and “medical staff and issues related to them” have the lowest average score of patient safety culture, and in the study of Macedo et al. The occurrence of errors (61%) is allocated (6). In the studies conducted by Tamazoni et al. (2015) in Brazil (18) and Arshadhi Bostan Abad et al. (2015) in Iran (4), the most negative response of nurses was related to non-punitive response in case of errors. Unlike the present study, in the next study by Salavati et al., the non-punitive response to the error had the highest safety culture score (27).

The obtained results can be due to the fact that nurses are facing problems in recognizing the importance of issues related to patient safety, and the presence of the traditional culture of blame and punishment is still tangible in the management method and show the need to change the current culture. Because blaming people leads to missed opportunities to learn from errors, better train care providers about how they operate and be alert to situations where the probability of errors is high, and on the other hand, prevents the creation of well-functioning processes to prevent from committing similar errors in the future. One of the biggest obstacles to preventing errors is punishing and blaming people for their mistakes(4).

## Conclusion

According to the fact that in the investigation of the cultural status of safety in this study, most of the nurses had an average level of safety culture, and considering the significant cultural importance of patient safety and its impact on providing safe care to children and preventing errors, the design, and implementation of the program Educational activities in the context of in-service continuous training courses, in the context of increasing the status of patient safety culture and improving the quality of nursing services and care, as well as to reduce the occurrence of errors and improve the safety level of sick children. It is suggested that similar studies in this field investigate the educational method to improve the cultural status of nurses working in children’s wards.

### How might this information affect nursing practice

Pediatric nurses can play an active role in patient safety. By relying on the weak points seen in this study and educating details in patient safety during patient care to nurses and nursing students; it is possible to increase the patient safety culture in them in order to improve the quality of nursing care and reduce the occurrence of errors and improve the safety level in pediatrics. Health center managers should create conditions for nurses to report errors without fear of punishment so that they can identify the root cause of the error and prevent the same error from occurring again in the future. They can help increase patients’ safety culture and consequently maintain and improve their health by making decisions with a non-punitive approach and using supportive measures.

### Limitation

One of the limitations of this study was the use of a self-report questionnaire, which attempted to encourage the participants to adhere to the principle of honesty by assuring the nurses of the confidentiality of their names and identities. Also, the crowding and heavy workload in the children’s department affected the nurses’ concentration while answering the questions, so they tried to go to the hospital in multiple shifts to complete the questionnaires during the nurses’ quiet and rest time.

## Ethics committee approval

The Institutional Ethics Committee of Guilan University of Medical Sciences approved this study (approval number: IR.GUMS.REC.1400.213, date: 12.10.2023).

## Consent for publication

None of the authors declared

## Availability of data and materials

The data and materials that support the findings of this study are available from the corresponding author upon reasonable request.

## Author contributions

Z.M. and M.M. designed the overall study. Z.T.E. participated in the recruitment and data collection. E.K.L. analyzed the data. All authors read and approved the final manuscript.

## Acknowledgments

The authors are very grateful to the volunteers who participated in this study and the Guilan University of Medical Sciences, Rasht, Iran.

